# COVID symptoms, testing, shielding impact on patient reported outcomes and early vaccine responses in individuals with multiple myeloma

**DOI:** 10.1101/2021.05.20.21257379

**Authors:** Karthik Ramasamy, Ross Sadler, Sally Jeans, Sherin Varghese, Alison Turner, Jemma Larham, Nathanael Gray, Joe Barrett, Stella Bowcock, Gordon Cook, Chara Kyriakou, Dean Smith, Mark Drayson, Supratik Basu, Sally Moore, Sarah McDonald, Sarah Gooding, Muhammad K Javaid

## Abstract

**Objective:** Multiple myeloma (MM)-related morbidity has a profound effect on quality of life (QoL), and immune function, but few studies have prospectively examined the impact of COVID-19 pandemic and attendant vaccination on both immunity and QoL of patients with MM. We aimed to characterise these effects in a prospective cohort study.

**Design:** We initiated a prospective national cohort study of patients with MM from start of the second wave of SARS CoV-2 infections in December 2020 and resultant COVID lockdown in the United Kingdom. We assessed current myeloma status, history of COVID19 symptoms, testing and vaccination including response using the rudystudy.org platform. In addition, healthcare resource use, mental and social well being and loneliness (Lubben scale) from the start of the COVID-19 pandemic were assessed.

**Participants:** We report data from the first one hundred and nine adults with MM who completed the questionnaires and the first round of blood testing in the cohort.

**Results:** Five patients (4.5%) had COVID-19 infection confirmed by history and/or testing (Nucleocapsid antibody). Up to 98% of patients shielded completely or partially during both waves of the pandemic, with 18% of patients consequently changing antimyeloma therapy in the shielding period. Using the Lubben scale, 21/99 (21.2 %) reported social isolation. Using HADS scale 23.1% of patients reported symptoms of mild to moderate anxiety or mild to moderate depression during this period. Humoral immune response (spike ab) tested 3 weeks after first vaccination was detected in 17/28 (60%) patients.

**Conclusion:** Myeloma patients shielded during waves of the pandemic with significant change to therapy, low level natural COVID-19 infection (4%) and social isolation. Humoral response following the first dose of COVID-19 vaccine is lower than that reported in non-myeloma cohorts.

**What is already known on this topic:** Limited published data exist on the effect of the COVID-19 pandemic on myeloma patients. Post first vaccine response in myeloma patients has been reported in a small number of patients from two studies ranging from 25 % to 56%.

**What this study adds:** This study reports myeloma patients shielded during waves of the pandemic and demonstrates consequent significant social isolation and changes to therapy. Low level natural COVID-19 infection (4%) was noted in the study and humoral response following first dose of COVID-19 vaccine was lower than that reported in non-myeloma cohorts.

## INTRODUCTION

Annually over 6,000 patients are diagnosed with myeloma in the UK with an estimated prevalence of 25,000. Patients with myeloma have been shielded and self isolated since the start of the COVID-19 pandemic, due to the concern and subsequent reports of higher risk of severe COVID-19 disease and mortality^1^. In addition, guidelines have encouraged attenuation of myeloma therapeutics or switch to oral therapies, ostensibly to reduce hostpital foot-fall, facilitate shielding and potentially to limit further treatment-related immune suppression^2^. Myeloma requires ongoing immune suppressive chemotherapy, frequent medical visits and has previously demonstrated universally poor response to vaccinations3

SARS-CoV-2, the novel coronavirus, may be life-threatening for myeloma patients, and there remains limited evidence of how they respond to SARS-CoV-2 infection when they survive, particularly in those who have not been hospitalised and therefore not captured in reports to date. Prolonged viral shedding and delayed or negligible serconversion has been observed^3^. Despite self-isolation, a proportion of patients with myeloma will be exposed to SARS-CoV-2 infection.. The proportion of patients with asymptomatic SARS-CoV-2 infection is not known. Understanding the nature of this response is key to rationalizing care in myeloma patients. The inherent immunodeficiency that MM patients acquire is multifactorial. The pathophysiology of the disease result in plasma cell loss/dysfunction,subsequent hypogammaglobulinemia and impaired lymphocyte function, alongside the immune suppressant effects of malignant bone marrow infiltration and treatment. MM commonly affect the elderly population, a cohort that principally suffer with co-morbidities and immunesenescence, further increasing their risk of developing complications from infections^4^. Patients with haematological malignancies who contract influenza have been shown to be high risk of serious adverse complications, including secondary bacterial lower respiratory tract infections and death^5^.

Influenza vaccination studies report that myeloma patients often require repeated doses of vaccine to mount optimal antibody response^6^. The poor seroconversion results from established cellular and humoral immune dysfunction, exacerbated by active treatment and disease response status. Early reports of immune response to first dose of Pfizer COVID-19 vaccination is concerning, with 8/44 (18%) patients with blood cancer mounting an adequate immune response compared with 94% in healthy controls^7.^ The protective titre of antibodies required to prevent re-infection is unclear, as is the ability to protect patients from SARS-COV2 virus variants of concern (VOC). Moreover, the role of the cellular immune response to SARS-COV2 is crucial, and to date has not been assessed in MM patients. Cycles of shielding and self isolation cause considerable physical and psychological distress for myeloma patients. High mortality rates with COVID-19 coupled with anticipated poor COVID-19 vaccine responses will increase isolation periods and be detrimental to their myeloma care. The longer term impact for both myeloma control and physical and mental well-being is concerning.

In order to address these evidence gaps, all of which directly affect patient management decisions, both immediately and in future vaccine planning, we initiated a national web-based prospective study of adults with MM from December 2020 using patient co-developed measures of COVID symptoms, testing, vaccination, SARS COV-2 immunity acquired by infection or vaccination and QoL impact with standardised generic and disease specific patient reported outcome measures.

## METHODS

This is a prospective, observational study. The study is based on the existing RUDYstudy.org platform (LREC 14/SC/0126 & RUDY LREC 17/SC/0501), an established online rare disease platform with online dynamic consent and patient reported outcome assessments^8^.

### Recruitment

To ensure reaching our recruitment target rapidly, multiple pathways for recruitment were employed. The study is currently open to any UK resident to join online or via 30 hospitals, and is promoted by the Myeloma UK national patient group. In addition, the clinical team can either include the letter of invitation and study flyer with hospital appointment letters, and then offer consent to participants during that clinic visit, or direct them to the study website to consider registering. Participants were requested to identify their lead hospital clinician. Where the lead clinician was a known haematologist, participants were considered to have a confirmed MM diagnosis. Informed online dynamic consent was obtained for all participants.

### Assessments

The main assessments are web-based. Once consented, participants were asked to complete validated patient reported questionnaires Mental Health (Hospital Anxiety and Depression scale^9^). Currently, there are no validated questionnaires for COVID-19 symptoms, testing, vaccination or impact on wellbeing and healthcare resource use. A group of four adults with myeloma co-developed, together with clinicians with expertise in myeloma, a set of questions that covered these aspects, (Appendix). This RUDY COVID-19 consensus questionnaire was added to the existing online assessment forms on the RUDY platform. Participants were also invited to enter information about their myeloma diagnosis and current status. Chemotherapy treatment was coded as proteasome inhibitors - ixazomib, carfilzomib, bortezomib; immunomodulatory drugs - thalidomide, lenalidomide, pomalidomide; CD38 antibody – daratumumab, Isatuximab; and others – Belantamab, bendamustine, cyclophosphamide, dexamethasone, other steroids.

### Sample acquisition and handling

All patients were sent sampling details with prepaid postal sample return boxes. These additional blood samples were taken alongside their standard of care testing if possible to minimise additional visits to a healthcare setting. Standard information letters were sent to managing clinicians identified by participants (GPs and hospital consultants). Participants received email reminders to obtain these samples. Samples were centrally received, logged, processed and either distributed to collaborators for assays, or appropriately stored at the Immunology Lab, Churchill Hospital, Oxford.

### Serological testing

A serum sample was collected from each patient; this was tested for antibodies against SARS-CoV2 nucleocapsid (N) or spike (S) protein. SARS-CoV2 N protein antibodies were measured by turbidimetry (Abbott), with samples that produced values of >1.4 considered to be positive. SARS-CoV2 S protein antibodies were measured via 2 different methods; by turbidimetry (Abbott)(IgG serology only), with a cut-off value of 50 considered to be a positive result, and by ELISA (The Binding Site)(IgG, IgA, IgM serology as a combined result), where values greater than those for the calibrator sample were considered to be positive.

### Statistical analysis

Each assessment was scored according to published algorithms. Descriptive statistics included t-tests and ANOVA for parametric outcome measures, Spearman correlations, and Kruskall-Wallis tests for non-parametric outcomes. Categorical results were evaluated by Chi-Squared and Fisher’s exact tests when individual cell counts were less than 10. HADS questionnaires submitted from 5^th^ January were scored for mild, moderate and servere anxiety and depression^9^. Patient-reported chemotherapy details from January 5^th^ 2021 to March 2021 were included. For patients without a current chemotherapy entry, we used historical entries up to 4 months prior to study entry. SARS-CoV2 antibody response data were compared with published data from patients without myeloma with a history of COVID infection or vaccination. Multivariate logistic regression analyses were performed to identify independent predictors of quality of life, adjusting for sex, age and myeloma status. As a sensitivity analysis, models were repeated in those with confirmed vs. possible MM. Significance was determined as p< 0.05.

## RESULTS

One hundred and nine adults with myeloma completed the COVID-19 questionnaire with a returned blood sample between 5^th^ February and 29^th^ March 2021 when the UK was still in lockdown. In some cases, the questionnaire was not fully completed by the patient, meaning that for some analyses the sample number varies between 100-109.

### Baseline Characteristics and evidence of SARs-CoV-2 infection

Patient characteristics are listed in

**During** the waves of the pandemic, only a minortity of patient study participants (6%) reported major symptoms of COVID-19. In the patients who were tested for COVID-19 only one patient was PCR positive at time of study entry (**Table 2**). The majority of the testing was performed as part of a routine screen prior to planned anti myeloma therapy.

**Table 1.**
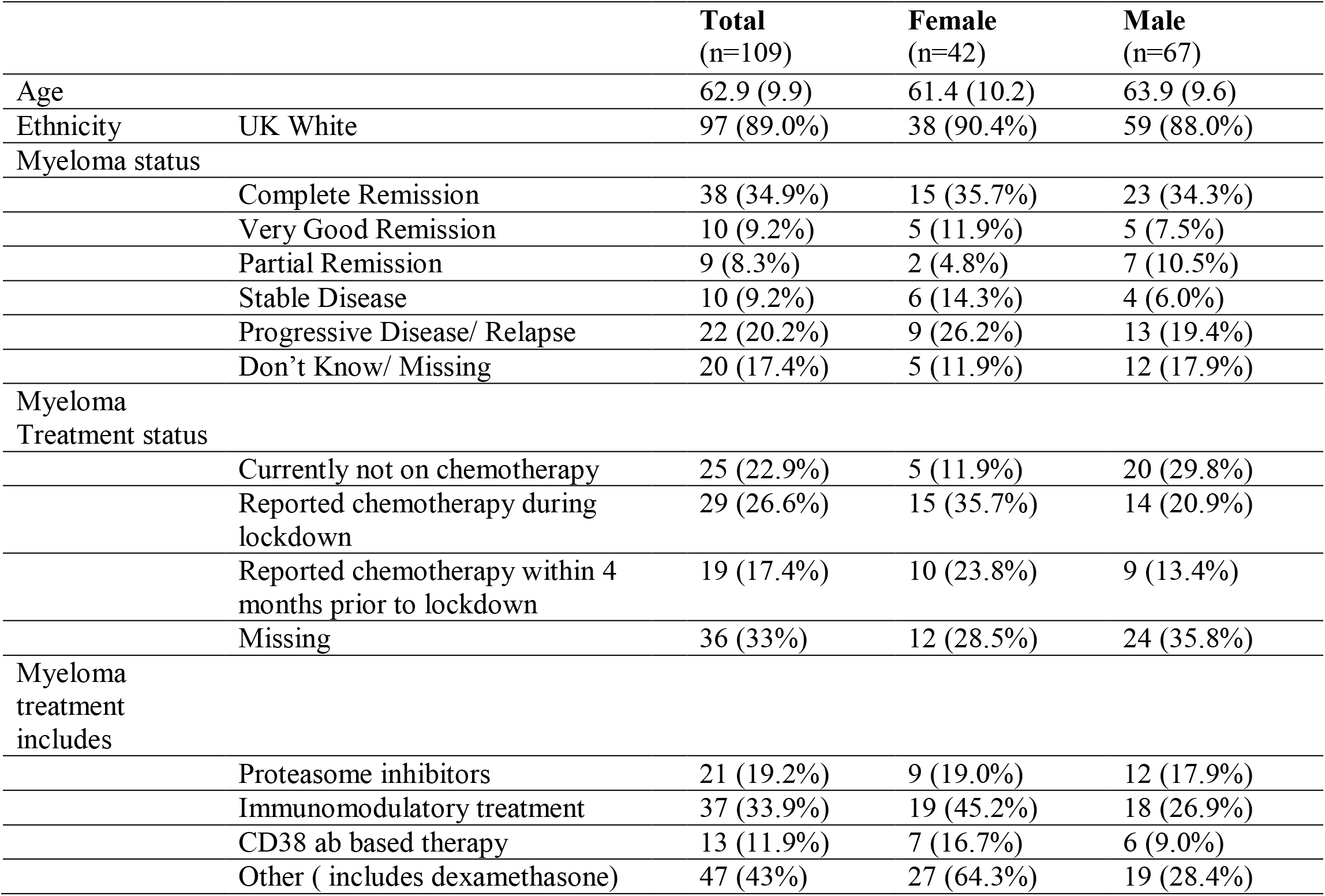
Patient characteristics of first 109 study participants who took part in the PREPARE study.

|  |  | **Total**<br>(n=109) | **Female**<br>(n=42) | **Male**<br>(n=67) |
| --- | --- | --- | --- | --- |
| Age |  | 62.9 (9.9) | 61.4 (10.2) | 63.9 (9.6) |
| Ethnicity | UK White | 97 (89.0%) | 38 (90.4%) | 59 (88.0%) |
| Myeloma status |  |  |  |  |
|  | Complete Remission | 38 (34.9%) | 15 (35.7%) | 23 (34.3%) |
|  | Very Good Remission | 10 (9.2%) | 5 (11.9%) | 5 (7.5%) |
|  | Partial Remission | 9 (8.3%) | 2 (4.8%) | 7 (10.5%) |
|  | Stable Disease | 10 (9.2%) | 6 (14.3%) | 4 (6.0%) |
|  | Progressive Disease/ Relapse | 22 (20.2%) | 9 (26.2%) | 13 (19.4%) |
|  | Don't Know/ Missing | 20 (17.4%) | 5 (11.9%) | 12 (17.9%) |
| Myeloma Treatment status |  |  |  |  |
|  | Currently not on chemotherapy | 25 (22.9%) | 5 (11.9%) | 20 (29.8%) |
|  | Reported chemotherapy during lockdown | 29 (26.6%) | 15 (35.7%) | 14 (20.9%) |
|  | Reported chemotherapy within 4 months prior to lockdown | 19 (17.4%) | 10 (23.8%) | 9 (13.4%) |
|  | Missing | 36 (33%) | 12 (28.5%) | 24 (35.8%) |
| Myeloma treatment includes |  |  |  |  |
|  | Proteasome inhibitors | 21 (19.2%) | 9 (19.0%) | 12 (17.9%) |
|  | Immunomodulatory treatment | 37 (33.9%) | 19 (45.2%) | 18 (26.9%) |
|  | CD38 ab based therapy | 13 (11.9%) | 7 (16.7%) | 6 (9.0%) |
|  | Other ( includes dexamethasone) | 47 (43%) | 27 (64.3%) | 19 (28.4%) |

**Table 2.** Characteristics of study participants who reported COVID symptoms and/or reported having PCR testing performed.

| Characteristics |  | Total<br>(n=109) | Male<br>(n= 67) | Female<br>(n=42) |
| --- | --- | --- | --- | --- |
| Potential COVID 19 symptoms | History of any major symptoms<sup>1</sup> | 8 (7.3%) | 7 (10.5%) | 1 (2.4%) |
|  | History of other potentially related symptoms<sup>2</sup> | 23 (21.1%) | 13 (19.4%) | 10 (23.8%) |
| Repeated potential COVID symptoms |  | 11 (10.1%) | 6 (9.0%) | 5 (11.9%) |
| Ever PCR tested |  | 64 (58.7%) | 44 (65.7%) | 20 (47.6%) |
| Reason for testing |  |  |  |  |
|  | Because of symptoms | 10 (16.1%) | 7 (15.9%) | 3 (15.0%) |
|  | As part of contact tracing | 2 (3.1%) | 1 (2.3%) | 1 (5.0%) |
|  | Screening before hospital treatments | 34 (53.1%) | 22 (50.0%) | 12 (60.0%) |
|  | Community testing | 4 (6.3%) | 3 (5.0%) | 1 (5.0%) |
|  | Research study | 12 (18.8%) | 9 (15.0%) | 3 (15.0%) |
|  | Missing reason | 2 (3.1%) | 2 (4.6%) | 0 |
| COVID Test results |  |  |  |  |
|  | Positive | 1 (1.6%) | 1 (2.3%) | 0 |
|  | Negative | 60 (93.4%) | 40 (90.9%) | 20 (100%) |
|  | Don't know | 2 (3.1%) | 2 (4.6%) | 0 |

Legend: <sup>1</sup>Major symptoms of high fever, cough and change in taste or smell. <sup>2</sup>Potential symptoms include fatigue, shortness of breath, loss of appetite, muscle pain, sore throat, headache, runny or stuffy nose, stomach toe rash, diarrhoea/digestive issues/upset stomach and nausea/ vomiting

### Changes in healthcare and wellbeing

Almost all patients were either partially or fully shielded during both waves of the pandemic Fewer patients fully shielded in the second wave in comparison with the first wave.. The primary reason for shielding was due to current or recent immunosuppressive therapy (**Table 3**). Respondants perception of the healthcare is shown in Table 4. When asked how NHS care had changed during the pandemic, 46/67 (68.7%) of women and 58/100 (58%) of men reported an improvement in care (Shaded in Blue **Figure 1**). A third of respondants did not feel that NHS care had improved during the pandemic. When asked about the type of consultation, 46.5% of participants preferred remote consultation over face to face appointments **Figure 2**. Men were more likely to favour video consultation than women after adjusting for age (OR 1.78 (95%CI 1.03 – 3.08)). with 10% of recruited adults non UK white. All data provided are from patient completed online entries. Chemotherapy data has been cross validated in up to 50% of patients using chemotherapy database at centres. Although the majority of patients reported they were in remission, 20% of patients reported they had poorly controlled myeloma, either relapse or progressive disease. A proportion of patients (22%) were not on active therapy with the rest of the patients on active therapy. Where reported, 11% were on antibody therapy, 33 % were on immunomodulatory drugs, 18 % were on proteasome inhibitors and 42 % were on other therapy.

**Table 3.** Level of Shielding and reasoning reported by participants in the PREPARE trial.

|  |  | **Total**<br>(n=102) | **Male**<br>(n=65) | **Female**<br>(n=37) |
| --- | --- | --- | --- | --- |
| Shielded during first lockdown |  | 101 (99%) | 65 (100%) | 36 (97.3%) |
| Shielding level during first lockdown |  |  |  |  |
|  | Complete | 65 (63.7%) | 39 (60.0%) | 26 (70.3%) |
|  | Partial | 47 (46.1%) | 34 (52.3%) | 13 (35.1%) |
|  | None | 2 (2.0%) | 1 (1.5%) | 1 (2.7%) |
| Current shielding level<sup>1</sup> |  |  |  |  |
|  | Complete | 41 (40.2%) | 24 (36.9%) | 17 (45.9%) |
|  | Partial | 63 (61.8%) | 42 (64.6%) | 21 (56.8%) |
|  | None | 1 (1.0%) | 1 (1.5%) | 0 |
| Reasons for shielding |  |  |  |  |
|  | Post-transplant | 11 (10.8%) | 7 (10.8%) | 4 (10.8%) |
|  | On steroids | 33 (32.4%) | 19 (29.2%) | 14 (37.8%) |
|  | Chemotherapy | 55 (53.9%) | 31 (47.7%) | 24 (64.9%) |
|  | Radiotherapy | 0 | 0 | 0 |
|  | Other treatments | 7 (6.9%) | 5 (7.7%) | 2 (5.4%) |
|  | Other reasons | 38 (37.3%) | 27 (41.5%) | 11 (29.7%) |

Legend: <sup>1</sup>Respondents were permitted to select more than one shielding level and reason for shielding

**Table 4.** Participant reported impact on healthcare resources due to COVID 19 in the PREPARE trial.

|  |  | **Total**<br>(n=101) | **Male**<br>(n=66) | **Female**<br>(n=35) |
| --- | --- | --- | --- | --- |
| Participant initiated |  |  |  |  |
|  | Cancel or postpone appointment | 7 (6.9%) | 2 (3.0%) | 5 (14.3%) |
|  | Change therapy to avoid hospital attendance | 18 (17.8%) | 12 (18.2%) | 6 (17.1%) |
| Hospital initiated change to appointment |  |  |  |  |
|  | Cancel or postponed | 22 (21.8%) | 11 (16.7%) | 11 (31.4%) |
|  | Change to phone call | 15 (14.9%) | 10 (15.2%) | 5 (14.3%) |
|  | Change to video call | 9 (8.9%) | 7 (10.6%) | 2 (5.7%) |
|  | Moved to another hospital | 4 (4.0%) | 3 (4.5%) | 1 (2.9%) |
| Therapy unavailable |  | 3 (3.0%) | 3 (4.5%) | 0 |
| Procedure postponed or cancelled |  |  |  |  |
|  | Surgical | 3 (3.0%) | 2 (3.0%) | 1 (2.9%) |
|  | Imaging | 2 (2.0%) | 1 (1.5%) | 1 (2.9%) |
|  | Day unit therapy | 28 (27.7%) | 19 (28.8%) | 9 (25.7%) |
|  | Allied health professional | 5 (5.0%) | 4 (6.1%) | 1 (2.9%) |
| Difficulty reaching speciality team for an urgent concern |  | 2 (2.0%) | 2 (3.0%) | 0 |

**Table 5.**
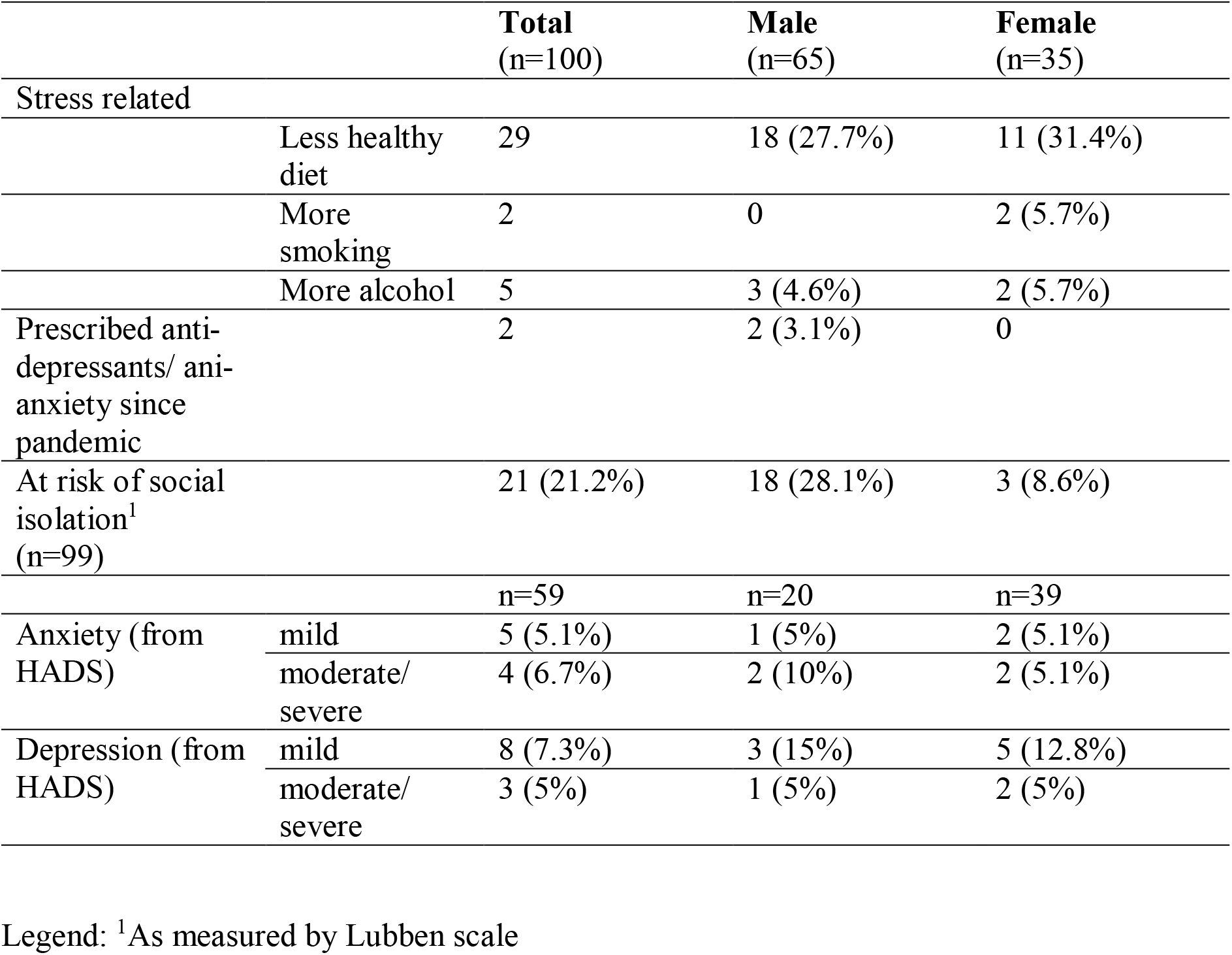
Participant reported impact on mental and social wellbeing due to COVID 19 in the PREPARE trial.

|  |  | **Total**<br>(n=100) | **Male**<br>(n=65) | **Female**<br>(n=35) |
| --- | --- | --- | --- | --- |
| Stress related |  |  |  |  |
|  | Less healthy diet | 29 | 18 (27.7%) | 11 (31.4%) |
|  | More smoking | 2 | 0 | 2 (5.7%) |
|  | More alcohol | 5 | 3 (4.6%) | 2 (5.7%) |
| Prescribed anti-depressants/ anti-anxiety since pandemic |  | 2 | 2 (3.1%) | 0 |
| At risk of social isolation<sup>1</sup><br>(n=99) |  | 21 (21.2%) | 18 (28.1%) | 3 (8.6%) |
|  |  | n=59 | n=20 | n=39 |
| Anxiety (from HADS) | mild | 5 (5.1%) | 1 (5%) | 2 (5.1%) |
|  | moderate/severe | 4 (6.7%) | 2 (10%) | 2 (5.1%) |
| Depression (from HADS) | mild | 8 (7.3%) | 3 (15%) | 5 (12.8%) |
|  | moderate/severe | 3 (5%) | 1 (5%) | 2 (5%) |

Legend: <sup>1</sup>As measured by Lubben scale

**Table 6.**
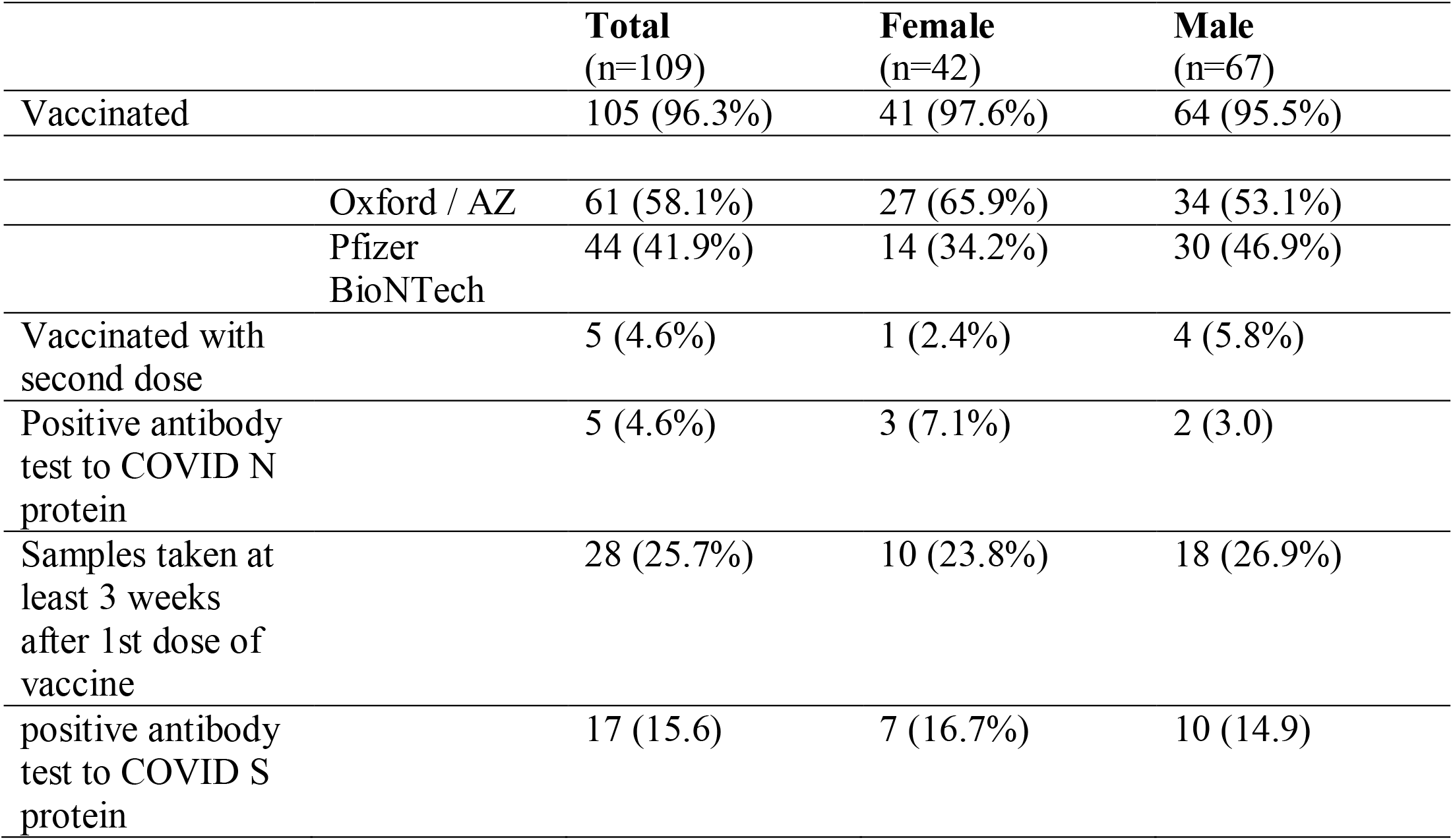
Vaccination status for 109 study participants in the PREPARE trial and their antibody response.

**Figure 1.**
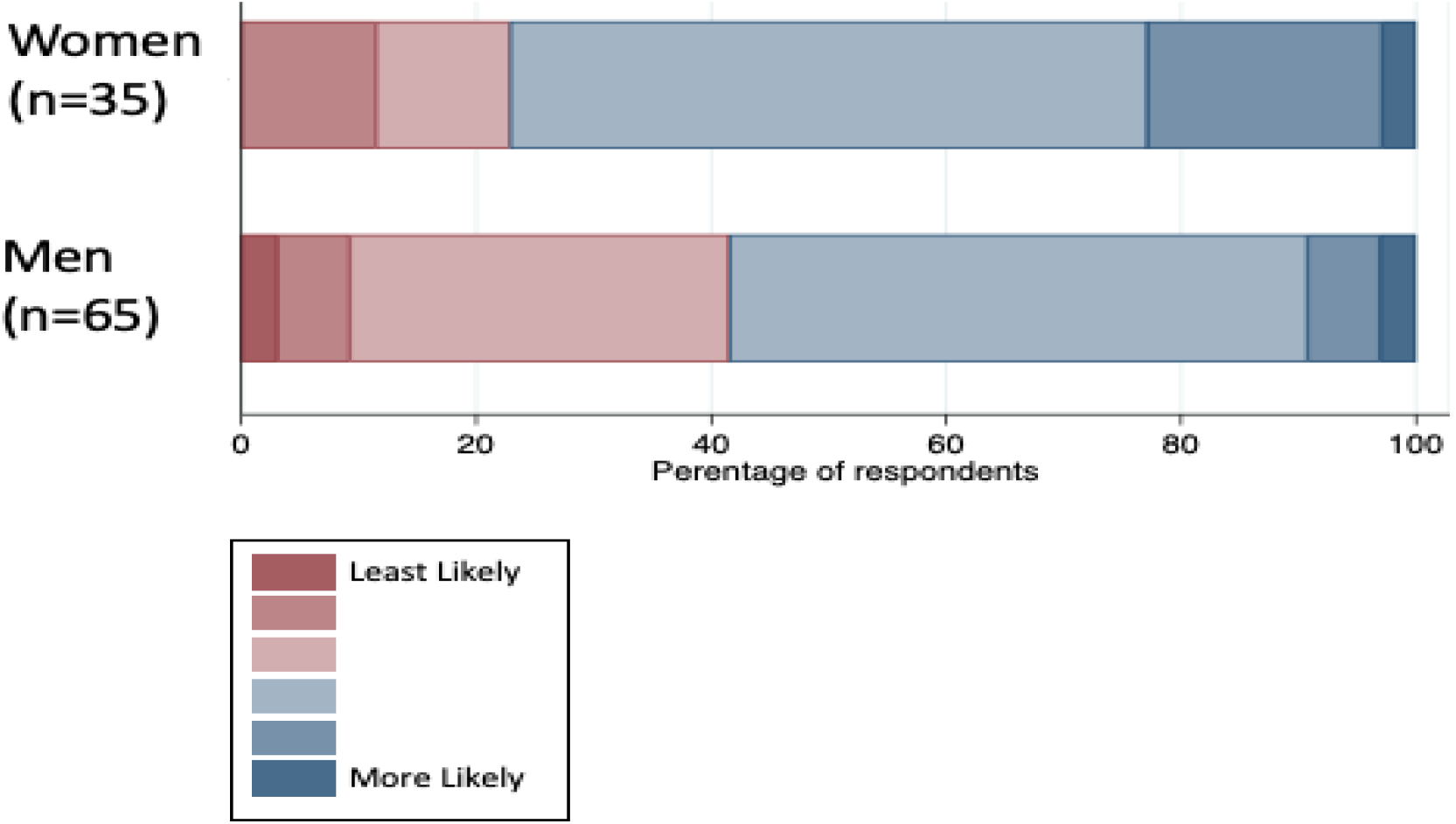
Participants’ opinion of change in quality of care received from the NHS during lockdown in comparison to prior NHS care. Questions and preferences in the RUDY COVID-19 questionnaire enclosed in the appendix.

**Figure 2.**
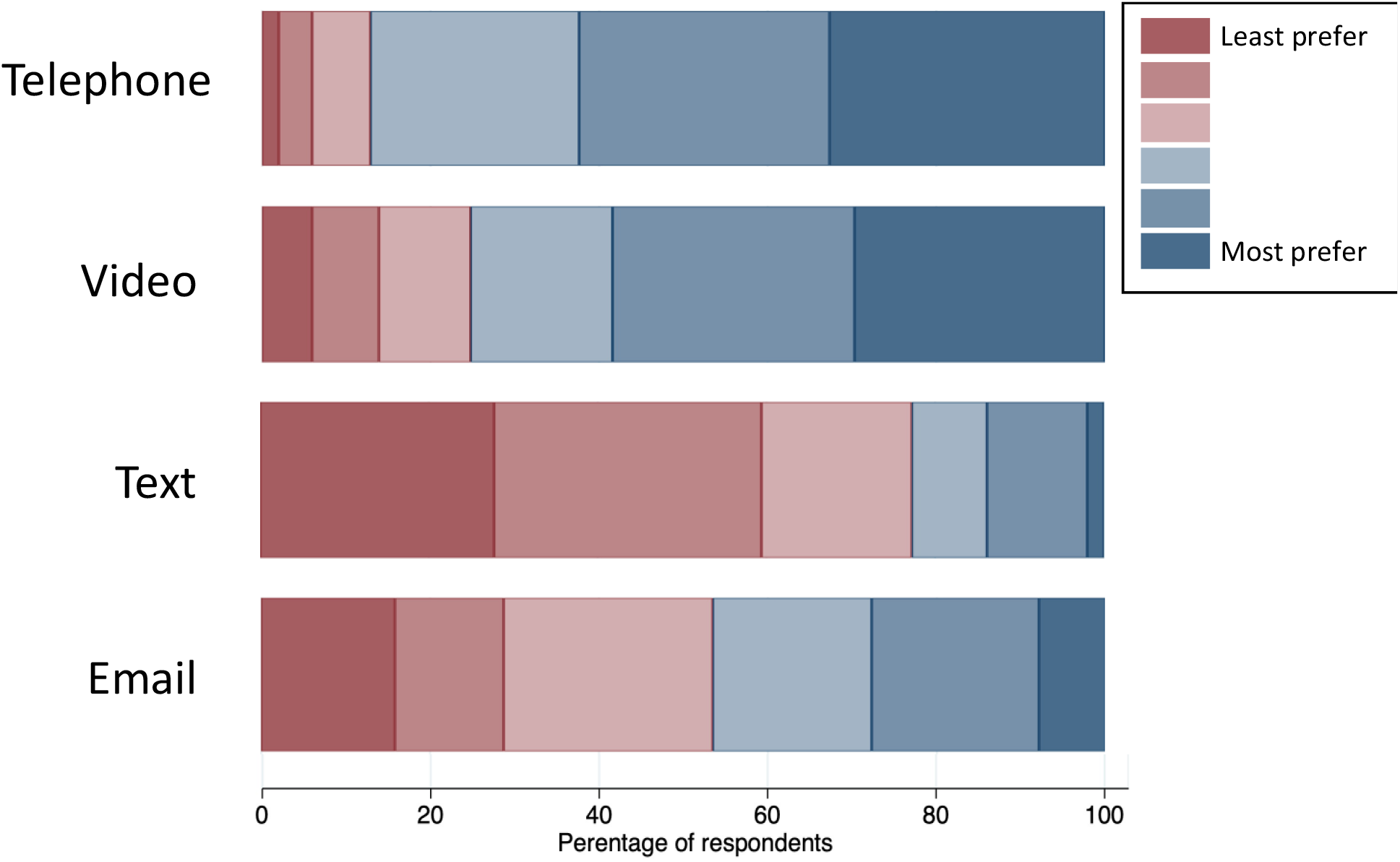
The above figure illustrates participant reported preferences for remote appointments during the pandemic. Questions and preferences in the RUDY COVID-19 questionnaire enclosed in the appendix.

During the waves of the pandemic, only a minortity of patient study participants (6%) reported major symptoms of COVID-19. In the patients who were tested for COVID-19 only one patient was PCR positive at time of study entry (**Table 2**). The majority of the testing was performed as part of a routine screen prior to planned anti myeloma therapy.

### Changes in healthcare and wellbeing

Almost all patients were either partially or fully shielded during both waves of the pandemic Fewer patients fully shielded in the second wave in comparison with the first wave.. The primary reason for shielding was due to current or recent immunosuppressive therapy (**Table 3**). Respondants perception of the healthcare is shown in Table 4. When asked how NHS care had changed during the pandemic, 46/67 (68.7%) of women and 58/100 (58%) of men reported an improvement in care (Shaded in Blue **Figure 1**). A third of respondants did not feel that NHS care had improved during the pandemic. When asked about the type of consultation, 46.5% of participants preferred remote consultation over face to face appointments **Figure 2**. Men were more likely to favour video consultation than women after adjusting for age (OR 1.78 (95%CI 1.03 – 3.08)). Almost a third of respondents identified with making less healthy diet choices during the pandemic but few reported increases in alcohol, smoking or medical treatment for anxiety or depression. One in 5 respondents scored ‘at risk of social isolation’, with no differences by age but men significantly more likely to be at risk than women (p=0.027). The reported impact of the pandemic on lifestyle and social activities varied considerably between participants (**Figure 3**). The majority respondents identified greater connection with friends and family, less social pressure, better finances, easier wearing of masks and informative reading or watching of the news. There were no differences by gender except for outdoor activities, which were less likely in men than women (47.7% vs 25.7% respectively, p=0.03).

**Figure 3.**
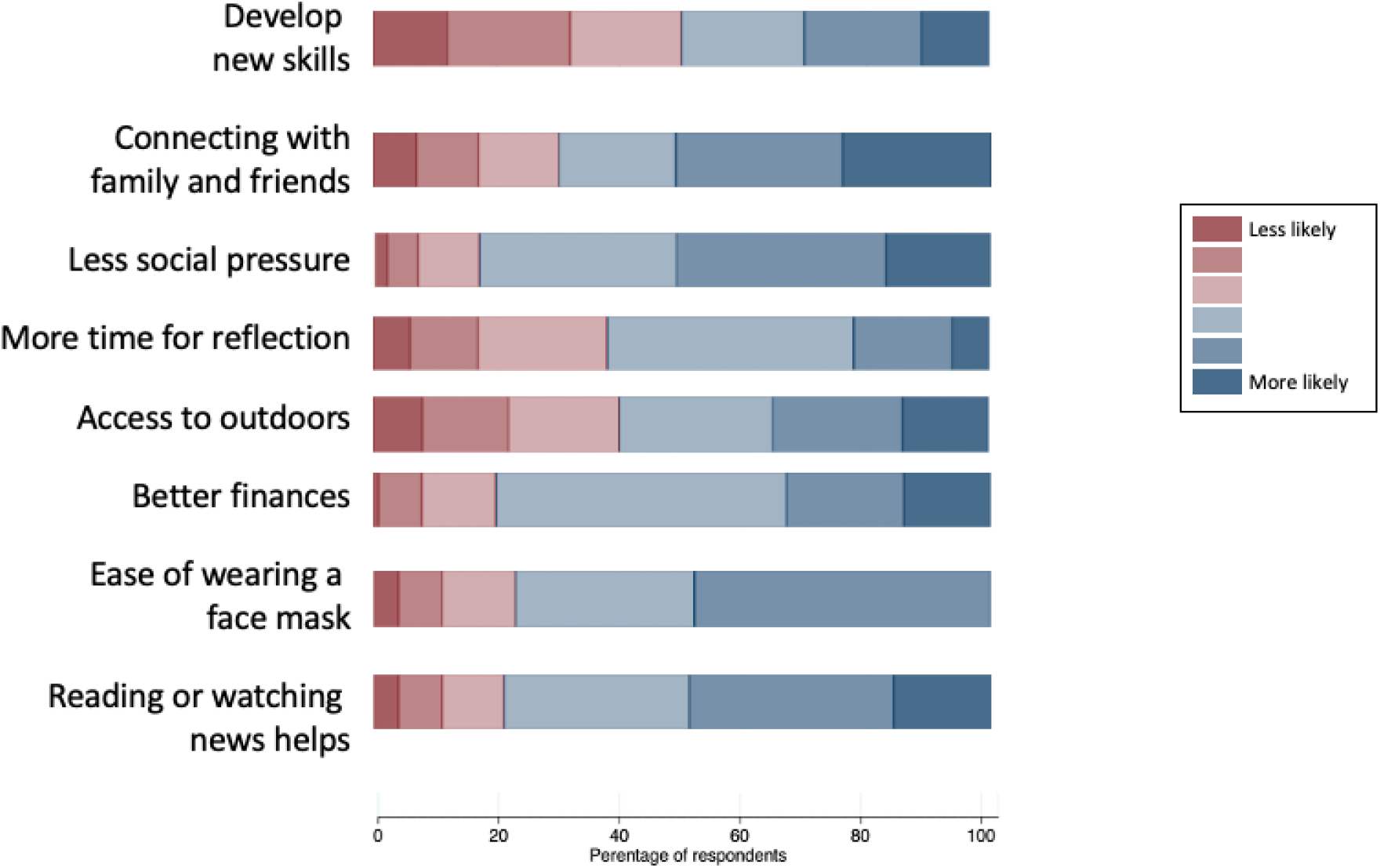
Participant reported impact^1^ on mental and social wellbeing due to COVID in the PREPARE trial. Questions and preferences in the RUDY COVID-19 questionnaire enclosed in the appendix. Legend: ^1^Stacked bar chart for how participants felt about a list of pre-defined activities during the last 4 weeks of lockdown.

### Humoral response to First dose of COVID-19 vaccination

2 patients samples were not suitable for serological analysis at the time of reporting. Out of the remaining 107 patients, 5 were found to have positive levels of N protein antibodies, indicating that they had suffered a natural infection during the pandemic and successfully generated an immune response against it. Only one of these patients had history of a PCR positive result known at study entry with the remaining patients having had asymptomatic infections. There were no instances of history of prior PCR-positive results but negative N protein antibodies. When looking at the timing of samples that were received from these patients, approximately 25% of them were taken from the patient >3 weeks post their 1^st^ dose of vaccine (range 21-68 days). 17/28 (60%) of these patients showed positive S protein antibodies, with 13 of these patients showing no evidence of natural infection with COVID and therefore indicating that they had produced a measurable response to the 1^st^ dose of their respective vaccine. 50% of patients who received Astra-Zeneca/Oxford vaccination and 44% of patients who received Pfizer vaccination produced a successful response > 3 weeks post 1^st^ dose (sample n = 14 (AZ) vs 9 (P)). Correlation with other factors such as age, disease status and therapy were not assessed due to smaller numbers.

## DISCUSSION

Myeloma patients have stayed either partially or fully shielded during the waves of the COVID-19 pandemic. This has resulted in significant change in their healthcare provision received from the NHS, with drug treatment modifications to enable shielding and reduce immunosuppression in 18%. Surprisingly, the majority of patients feel their care has improved during the pandemic but a third of patients report poor care.. Patients continue to prefer face to face appointments in clinic rather than remote consultations. Up to 20% of patients are at risk of social isolation. In addition, patients experienced higher degrees of depression and anxiety compared to the pre pandemic state. Although only one patient had known prior infection, a few patients (4/109) have evidence of asymptomatic COVID-19 infection. All patients who have had a previous infection had a robust antibody response to the first dose of COVID-19 vaccine and 60% of patients have an optimal immune response to COVID-19 vaccination. There were no differences noted in humoral response after first dose of either Astrazeneca (Adenoviral vector) or Pfizer mRNA based vaccination.

### Strengths and weaknesses of the study

Previous myeloma studies during the pandemic have primarily focussed on outcomes of COVID-19 infection in hospitalised myeloma patients. This is the first prospective study to report on history of COVID19 symptoms, testing, healthcare resource impact, and mental and social well being in the shielded population during both waves of the COVID-19 pandemic from a community based sample. This study highlights the effects of long term shielding / self -isolation, and changes in heathcare provision, have had on social and mental well being of myeloma patients. The effects are comparable to other cancer patients with incurable disease and require ongoing healthcare support^13^. The study was planned and recruitment started during the pandemic, which made the study exclusively online. This has biased the demographic to a relatively well, computer literate and younger cohort of patients The median age of the population is younger (62.9 yrs) than the typical median age at diagnosis (69 yrs) as computer literacy enables participation in this study. This may also have under represented the proportion of patients experiencing social isolation and either anxiety or depression. Details on chemotherapy were patient reported and therefore not fully captured through the online portal; and work is ongoing to record them from individual centres. Our study design does not capture the profoundly immunosuppressed MM patients who may have succumbed to SARS-CoV-2 infection.

During the pandemic, large scale changes in healthcare were introduced to support shielding practices and to limit exposure to immunosuppressive chemotherapy. Consults were often remote through telephone or video calls and patients had only blood tests performed if absolutely needed to plan care. The findings of this study illustrate the variable experiences of patients with myeloma in terms of changes in hospital visits and therapies, such as day unit access. While a approximately a third reported a reduction in the quality of care from the NHS, reassuringly very few reported difficulties in contacting their speciality team for an urgent concern. When questioned about future types of consultation, nearly 50% most preferred telephone and video over text and email consultations.

This study has described concerning rates of social isolation. Social isolation has been one of the key issues affecting patients during COVID-19. COVID-19 can trigger anxiety and distress, which may even be of increased intensity especially in vulnerable patient groups such as oncology patients^10,13^. It was noted that patients with cancer reported higher level of isolation and even experienced emotions of guilt, due to dependence on family members^10,13^. Furthermore, after controlling for prior depression and other factors, it was loneliness due to social isolation during COVID-19 that increased the risk of depression especially for cancer of the breast, prostate, and blood ^11,^. Both loneliness and social distancing also increases risk of mortality in cancer patients, while feelings of uncertainty increased emotional distress^12,13^. Multiple sources have suggested the need for more frequent contact, via telephone or video consultation, with healthcare professionals to increase social support^14,15^. Issues regarding trust, isolation and worries about abandonment should also be proactively addressed^16^.

Just over 50% of respondants completed the HADS. Of the respondants, 6.7% reported moderate to severe anxiety, with higher rates in men than women. Similar rates of depression were seen with little differences between sexes. Quinn et al reported a higher degree of anxiety and depression (60%) using WHOQOL-BREF questionnaire during the first wave, in a snapshot international online survey of myeloma patients ^17^. This is much higher than reported in our study. The differences could be explained in differences in questionnaire, by adjustments over the one year period as well as a relative low level of transmission of COVID-19 between July 2020 and September 2020, and that patients were able to socialise with family and friends in this period.

The patients reported here, at the time of analysis, are yet to receive the second dose of COVID-19 vaccine, therefore both magnitude of humoral response reported and Tcell responses could change following second dose of the vaccine. However, these data suggest that where COVID vaccines are not widely available, offering single dosing or delaying the second dose in this patient group should be avoided.

### Comparison with existing literature

Our study is unique in narrating the journey of myeloma patients through both waves of the COVID-19 pandemic. Other studies have primarily focussed on immune responses in myeloma patients following first dose of COVID-19 vaccination. Terpos et al report that 28.2% of patients generated neutralising antibody titres following a first dose of Pfizer COVID-19 vaccine and Bird et al report a 56% antibody response to a first dose of vaccine ^18,19^. We have noted antibody responses in 60% of patients using the same antibody platform as Bird et al ^19^. Smaller patients numbers explain differences in early humoral immune response between these studies. An older age cohort could explain the lower proportion of patients with adequate antibody titres in the study by Terpos et al^18^. Higher humoral immune responses has been reported in a community study with a comparable age cohort following the first dose of either Pfizer or Astra Zeneca vaccine^20^. In a snapshot international online survey of myeloma patients, Quinn et al found 60% respondents reported anxiety and depression during the first wave ^17^. This is much higher than reported in our study. The differences could be explained by differences in questionnaires used, adjustments over the one year period, as well as a relative low level of transmission of COVID-19 between July 2020 and September 2020, meaning that patients were able to socialise with family and friends in the period before our study commenced.

### Implications for clinicians and policymakers

This study captures the challenges in managing immunosuppressed patients who are at a higher risk of morbidity and mortality from multiple waves of the COVID-19 pandemic. Our data supports early reports that suggest myeloma patients have relatively poorer vaccine immune response in comparison with their age-matched peers^20,21)^, which fuels concerns that such patients should continue to shield despite lockdowns easing. Further waves of pandemic and poor immunity to vaccination will continue to harm the social and mental wellbeing of these patients. These data suggest that urgent measures should be taken to suitably modify healthcare provision and provide psychological support for these patients. Measures to improve COVID-19 immunity such as additional booster vaccination or passive antibody trials should be prirotitised for this patient population to enable their full release from shielding behaviours, which would improve their mental wellbeing and their ability to stay on anti-myeloma therapy, which is vital to keep their terminal disease under control for as long as possible. *Longer term studies will assess whether myeloma survival outcomes have been negatively impacted by the therapeutic modifications which were reported in nearly 1 in 5 cases*. Social isolation has been one of the key issues affecting patients during COVID-19. COVID-19 can trigger anxiety and distress, which may even be of increased intensity especially in vulnerable patient groups such as oncology patients^10,13^. Both loneliness and social distancing also increases risk of mortality in cancer patients, while feelings of uncertainty increased emotional distress^12,13^. Multiple sources have suggested the need for more frequent contact, via telephone or video consultation, with healthcare professionals to increase social support^14,15^. Issues regarding trust, isolation and worries about abandonment should also be proactively addressed^16^.

### Unanswered questions and future research

Further follow up serology and cell-based testing, after administration of a second dose of vaccine, to measure both antibody response and T cell response is required. Our study has captured primarily younger, reasonably well and computer literate patients. the effects on social and mental well being as well as vaccine response could be poorer in an older cohort of myeloma patients. We plan to continue to recruit patients more representative of typical myeloma population cohort with a median age of 70.

## Supporting information

HRA approval

Ethics permission

## Data Availability

Patients have consented to anonymised data sharing for research and plublication purposes.

## Acknowledgements

Blood cancer UK, Myeloma UK patient panel, RUDY patient panel

## Contributor and Guarantor Information

All listed authors made substantial contributions to the conception or design of the work; or the acquisition, analysis, or interpretation of data for the work; and drafting the work or revising it critically for important intellectual content; final approval of the version to be published; and agreement to be accountable for all aspects of the work in ensuring that questions related to the accuracy or integrity of any part of the work are appropriately investigated and resolved. KR is the guarantor and accepts full responsibility for the work and/or the conduct of the study, had access to the data, and controlled the decision to publish.

## Copyright/Licence for Publication

The Corresponding Author has the right to grant on behalf of all authors and does grant on behalf of all authors, an exclusive licence (or non-exclusive for government employees) on a worldwide basis to the BMJ Publishing Group Ltd to permit this article (if accepted) to be published in BMJ editions and any other BMJPGL products and sub-licences such use and exploit all subsidiary rights, as set out in our licence.

## Data sharing statement

Patients have consented to anonymised data sharing for research and plublication purposes.

## Public and patient involvement statement

RUDY patients forum, Oxford Blood Group, Myeloma UK patient research panel were all consulted and feedback secured on study design, questionairre

## Ethics approval

The study is based on the existing RUDYstudy.org platform (LREC 14/SC/0126 & RUDY LREC 17/SC/0501), an established online rare disease platform with online dynamic consent and patient reported outcome assessments. IRAS no: 213780, RUDY study Minor amendment 3, HRA approval 28^th^ January 2021.

## Transparency declaration

Karthik Ramasamy, the lead author (and the manuscript’s guarantor), affirms that the manuscript is a honest, accurate, and transparent account of the study being reported; that no important aspects of the study have been omitted; and that any discrepancies from the study as planned (and, if relevant, registered) have been explained.

## Dissemination declaration

Not applicable

**Appendix 1:** RUDY COVID-19 consensus questionnaire

