## Appendix 1 for "COVID symptoms, testing, shielding impact on patient reported outcomes and early vaccine responses in individuals with multiple myeloma"

The Botnar Research Centre
The University of Oxford
Nuffield Orthopaedic Centre,
Oxford, OX3 7LD, United Kingdom
Study phone number: 07775541615

**COVID FOLLOW UP QUESTIONNAIRE– INCOMPLETE / COMPLETE / SUBMITTED: .......**ERROR! BOOKMARK NOT DEFINED.

VACCINE .................................................. **ERROR! BOOKMARK NOT DEFINED.**
SYMPTOMS (I.E. PHYSICAL AND MENTAL COMPLAINTS) ........................ **ERROR! BOOKMARK NOT DEFINED.**
LONG COVID QUESTIONNAIRE FOR THOSE WHO HAVE RECOVERED FROM COVID - ............. **ERROR! BOOKMARK NOT DEFINED.**
LEVEL OF SHIELDING.......................................... **ERROR! BOOKMARK NOT DEFINED.**
ACCESS TO MEDICAL CARE/HOW WERE YOUR HEALTHCARE NEEDS MET IN THE LAST 4 WEEKS ........**ERROR! BOOKMARK NOT DEFINED.**
THIS SECTION CONCERNS YOUR MENTAL & EMOTIONAL HEALTH ................... **ERROR! BOOKMARK NOT DEFINED.**
*Lubben Questionnaire: The next series of questions will help us get a picture of how supported you feel with respect to your social connections with family and friends. ..................**Error! Bookmark not defined.***

### RUDY COVID Questionnaire

#### GENERAL INTRODUCTION

The research team at RUDY want to understand and compare the experience of living through the first and second lockdown due to COVID-19 in RUDY participants including potential vaccine use. We also ask those with confirmed COVID-19, if there are any ongoing health concerns, now described as 'Long COVID syndrome'. We may use your information for research to improve how your care is delivered in the future.

#### COVID BASELINE QUESTIONNAIRE – incomplete / complete / submitted

##### Vaccine

Have you had a COVID vaccine?

a. Yes b. No

i. If Yes,

1. Do you know the name of the vaccine:
a. Pfizer BioNTech
b. Moderna vaccine
c. Oxford / AstraZeneca
d. Valneva
e. GSK/ Sanofi Novavax
f. Other
g. Don't know

2. Date of first injection:
3. Were you expected a second dose? Yes, No, Don't Know
   If yes > Have you had the second dose > Not yet, Yes
   If yes Date of second dose

##### Possible COVID Symptoms

Did you have any of these symptoms that could be due to COVID 19 from January 2020 onward?

A) Which Major Complaints

a. A high temperature (37.8$^0$C or above)
b. New and continuous cough
c. A loss of, or change, in your normal sense of taste or smell (anosmia)

B) Other symptoms that could be caused by COVID-19

a. Rash on the toes, also known as COVID toe
b. Shortness of breath
c. Fatigue
d. Loss of appetite
e. Muscle pain
f. Sore throat
g. Headache
h. Runny or stuffy nose
i. Diarrhoea/digestive issues/upset stomach
j. Nausea/vomiting

C) NEVER HAD THESE SYMPTOMS

IF yes> Have you had these any of these symptoms more than once? YES NO

Please enter date(s) any symptoms first started. If you can't remember the day or month you can leave them blank. dd/mm/yy <option to add another event>

Have you had a test (s) for COVID-19

a. No
b. Yes

*(Separate link to be created if the answer is yes)*

What was the result of your coronavirus test(s) select all that apply?

1. Positive—it showed I had coronavirus -
2. Negative---it showed I did not have coronavirus
3. Inconclusive
4. Don’t know

Why did you have the test(s)?

1. Because of your physical complaints (symptoms)
2. As part of contact tracing

3. As part of a screening before hospital treatments (e.g. operations or chemotherapy)
4. As part of community testing
5. Other reasons – please describe <need to add a free text box>

<If positive test>

Have you had more than one positive test for COVID more than 4 weeks apart: yes / no

For each positive test please enter the date and treatment you received.

Date of test. If you can't remember the day or month you can leave them blank. dd/mm/yy

What treatment(s) did you have (select all that apply)?

a. no symptoms
b. managed symptoms at home
c. Had to phone 111 in England, Wales and Northern Ireland or NHS 24 in Scotland
d. Had to visit GP surgery
e. Had to have home visit
f. Had to visit A&E
g. Admitted to stay overnight in hospital (number of days admitted)
h. Needed Oxygen – via mask
i. Admitted to Intensive care (days in ITU and site)

<Option to add another positive test and treatment>

< *Long Covid Questionnaire should only be asked if patients say they have had a positive COVID test>*

#### Long Covid Questionnaire for Those Who Have had a diagnosis of Covid

Some patients experience symptoms even after recovering from COVID in what is now called a LONG COVID syndrome if they start during or after the COVID infection and continue for at least 12 weeks / 3 months without another cause being found.

If you have recovered from COVID, are you still experiencing any of the following (please tick all that apply):

☐ Fatigue that interferes with daily activities

☐ Muscle or body aches

☐ Shortness of breath or difficulty in breathing

☐ Difficulty concentrating or focusing

☐ Inability to exercise or be active

☐ Headache

☐ Difficulty sleeping

☐ Anxiety

☐ Memory problems

☐ Dizziness

☐ Other symptoms, please specify __________

☐ No ongoing symptoms __________

Overall, I feel that I have received enough support from the GP or hospital to manage the above conditions:

| 0 | 1 | 2 | 3 | 4 | 5 |
|---|---|---|---|---|---|
| Do not agree | | | | | Strongly agree |

#### Level Of Shielding

Have you ever received a letter from the NHS or Chief Medical Officer saying you have been identified as someone at risk of severe illness if you catch coronavirus, because you have an underlying disease or health condition?

1. Yes
2. No

What level of shielding were you doing during the FIRST lockdown (tick ALL that apply):

1. Complete shielding (only leaving the home for hospital emergencies)

2. Partial shielding (maintaining social distancing and wearing a face mask outside the home, essential visits such as groceries and hospital appointments only, limiting meeting visitors and family inside and outside the home
3. I was not shielding or taking special precautions

Are you CURRENTLY receiving treatment or taking medications that may affect your immune system? (Please tick all that apply)

1. Medication following an organ transplant/bone marrow treatment
2. Medicines such as steroid tablets that weaken the immune system
3. Targeted therapy or chemotherapy for cancer treatment
4. Radiotherapy for cancer treatment
5. Other treatment or medication that may affect the immune system
6. None of these

What level of shielding are you doing TODAY? (tick only ONE)

1. Complete shielding (only leaving the home for hospital emergencies)
2. Partial shielding (maintaining social distancing and wearing a face mask outside the home, essential visits such as groceries and hospital appointments only, limiting meeting visitors and family inside and outside the hom
3. I was not shielding or taking special precautions

#### Access To Medical Care/How Were Your Healthcare Needs Met

By understanding this impact, researchers will be able to advise on how to improve services going forward.

1. Have ***YOU*** *ever* cancelled or postponed an appointment or treatments due to COVID-19:
    a. Yes
    b. No

    If Yes, what was the reason? Tick all that apply:
        i. I was afraid to leave the house to get treatment

ii. I felt it was more important to avoid possible exposure to the virus at health care facilities
iii. Problems or concerns about getting to appointment or hospital
iv. I could not reach my healthcare provider to obtain reassurance that hospital/clinic attendance was safe
v. Other- please add

2. Have ***YOU*** *ever* had to switch or change any aspect of your current therapies to avoid hospital attendance in the COVID-19 pandemic:
    a. Yes
    b. No

3. Have ***YOU*** *ever* had to switch or change any aspect of your current therapies due to medication unavailability in the COVID-19 pandemic:
    a. Yes
    b. No

4. I had a medical appointment changed, postponed or cancelled by the ***HOSPITAL*** due to COVID: (select all that apply if you have had more than one appointment).
    a. No change to appointment
    b. Postponed
    c. Cancelled
    d. Changed to phone call
    e. Changed to video call
    f. Moved to a different hospital

5. I had a surgical procedure postponed or cancelled due to COVID by the ***HOSPITAL***:
    a. Yes
    b. No

6. I had a scan, such as an x-ray or Ct or MRI, postponed or cancelled due to COVID by the ***HOSPITAL***:
    a. Yes
    b. No

7. I had a day unit treatment such as an iv infusion delayed or cancelled due to COVID by the ***HOSPITAL***:
    a. Yes
    b. No

8. I had an appointment with an allied health care professional (e.g. OT, physio, nurse practitioner, hearing tests) postponed or cancelled due to COVID by the ***HOSPITAL***:
    a. Yes

    b. No

9. I had difficulty reaching my speciality team for an urgent concern that could be related to my rare disease (such as pain management) due to COVID:
    a. Yes
    b. No

10. I prefer a *hospital* telephone/video consultation to a face-to-face appointment:
    a. Yes
    b. No

11. Please rate your hospital telephone/video consultation from 0-5 (0 poor experience, 5 excellent) _________

12. Which forms of virtual support would you most prefer for ongoing management of your health condition at home (0 least prefer, 5 most prefer)
    a. Telephone calls

| 0 | 1 | 2 | 3 | 4 | 5 |
|---|---|---|---|---|---|
| Least prefer | | | | | Most prefer |

    b. Video consultations

| 0 | 1 | 2 | 3 | 4 | 5 |
|---|---|---|---|---|---|
| Least prefer | | | | | Most prefer |

    c. Text messages/SMS

| 0 | 1 | 2 | 3 | 4 | 5 |
|---|---|---|---|---|---|
| Least prefer | | | | | Most prefer |

    d. Email

| 0 | 1 | 2 | 3 | 4 | 5 |
|---|---|---|---|---|---|
| Least prefer | | | | | Most prefer |

13. I had to attend A/E during the lockdown for a non-COVID problem

a. Yes
b. No

#### This Section Concerns Your Mental & Emotional Health

We appreciate the COVID pandemic has affect people in different ways. How have you felt about the following since the pandemic started

2. : Should be 0 – 5

Developing new skills or hobbies e.g. baking. (LESS likely > MORE Likely)

Connecting with family and friends (LESS likely > MORE Likely)

Care from the NHS during this time (WORSE > BETTER)

Social commitments (MORE pressure > LESS pressure)

Time for Spiritual and intellectual reflections. (REDUCED > INCREASED)

Access to the outdoors (REDUCED > INCREASED)

Money and finances (WORSE > BETTER)

Wearing of a face mask outside the home (WITH DIFFICULTY > EASY )

Reading or watching the news (NOT HELPFUL > HELPFUL)

Going to Accident and Emergency if you were to have an urgent problem. (LESS Confident > MORE Confident)

I have been eating LESS healthily because of comfort eating due to stress

☐ Yes ☐ No

I have been smoking MORE than usual because of stress.

☐ Yes. ☐ No ☐ N/A

I have been consuming MORE alcohol than usual because of stress

☐ Yes ☐ No ☐ N/A

I have been prescribed anti-depressants or anti-anxiety drugs since the pandemic started

☐ Yes ☐ No ☐ N/A

Please describe other ways the pandemic has affected you:

Lubben Questionnaire: The next series of questions will help us get a picture of how supported you feel with respect to your social connections with family and friends.

FAMILY: Considering the people to whom you are related by birth, marriage, adoption, etc

1. How many relatives do you see or hear from at least once a month?
   0 = none 1 = one 2 = two 3 = three or four 4 = five to eight 5 = nine or more
2. How many relatives do you feel at ease with that you can talk about private matters?
   0 = none 1 = one 2 = two 3 = three or four 4 = five to eight 5 = nine or more
3. How many relatives do you feel close to such that you could call on them for help?
   0 = none 1 = one 2 = two 3 = three or four 4 = five to eight 5 = nine or more

FRIENDSHIPS: Considering all of your friends including those who live in your neighbourhood

4. How many of your friends do you see or hear from at least once a month?
   0 = none 1 = one 2 = two 3 = three or four 4 = five to eight 5 = nine or more
5. How many friends do you feel at ease with that you can talk about private matters?
   0 = none 1 = one 2 = two 3 = three or four 4 = five to eight 5 = nine or more
6. How many friends do you feel close to such that you could call on them for help?
   0 = none 1 = one 2 = two 3 = three or four 4 = five to eight 5 = nine or more
