## Supplementary material for "COVID symptoms, testing, shielding impact on patient reported outcomes and early vaccine responses in individuals with multiple myeloma": HRA approval

**From:** <> **Date:** Thursday, 10 December 2020 at 10:21 **To:** Kassim Javaid <>, <> **Subject:** IRAS Project ID 213780. HRA and HCRW Approval for the Amendment

Dear Dr Javaid,

| | |
|---|---|
| **IRAS Project ID:** | 213780 |
| **Short Study Title:** | Rare and Undiagnosed Diseases Study (RUDY) |
| **Amendment No./Sponsor Ref:** | Substantial amendment 3 |
| **Amendment Date:** | 04 December 2020 |
| **Amendment Type:** | Substantial Non-CTIMP |

I am pleased to confirm **HRA and HCRW Approval** for the above referenced amendment.

You should implement this amendment at NHS organisations in England and Wales, in line with the guidance in the amendment tool.

**User Feedback**

The Health Research Authority is continually striving to provide a high quality service to all applicants and sponsors. You are invited to give your view of the service you have received and the application procedure. If you wish to make your views known please use the feedback form available on the HRA website: http://www.hra.nhs.uk/about-the-hra/governance/quality-assurance/.

Please contact for any queries relating to the assessment of this amendment.

Kind regards

**Ms Sarah Ferry**

**Health Research Authority**

Ground Floor | Skipton House | 80 London Road | London | SE1 6LH
**E.**
**W.** www.hra.nhs.uk
Sign up to receive our newsletter HRA Latest.
