## Supplementary material for "COVID symptoms, testing, shielding impact on patient reported outcomes and early vaccine responses in individuals with multiple myeloma": Ethics permission

**South Central - Berkshire B Research Ethics Committee**
The Old Chapel
Royal Standard Place
Nottingham
NG1 6FS

**Please note: This is the favourable opinion of the REC only and does not allow the amendment to be implemented at NHS sites in England until the outcome of the HRA assessment has been confirmed.**

10 December 2020

Heather House
Oxford University Hospitals NHS Foundation Trust
OUH R&D, Joint Research Office
Block 60, Churchill Hospital
OX3 7LE

Dear Heather House

| | |
|---|---|
| **Study title:** | **Rare and Undiagnosed Diseases Study (RUDY)** |
| **REC reference:** | **17/SC/0501** |
| **Protocol number:** | **1** |
| **Amendment number:** | **Substantial amendment 3** |
| **Amendment date:** | **04 December 2020** |
| **IRAS project ID:** | **213780** |

The above amendment was reviewed by the Sub-Committee in correspondence.

**Ethical opinion**

The members of the Committee taking part in the review gave a favourable ethical opinion of the amendment on the basis described in the notice of amendment form and supporting documentation.

**Approved documents**

The documents reviewed and approved at the meeting were:

| *Document* | *Version* | *Date* |
|---|---|---|
| Completed Amendment Tool [Amendment Tool] | | 07 December 2020 |
| Copies of materials calling attention of potential participants to the research [RUDY-prepare_leaflet] | 1 | 01 December 2020 |
| Covering letter on headed paper [RUDY sub amendment 3 cover letter v1] | 1 | 01 December 2020 |
| Letters of invitation to participant [RUDY Letter for pt having blood taken v2(tracked)] | 2 | 08 December 2020 |
| Letters of invitation to participant [RUDY Letter for pt having blood taken v2] | 2 | 08 December 2020 |
| Letters of invitation to participant [RUDY BLOOD AND URINE PIS Adult] | 3 | 08 December 2020 |
| Other [RUDY-COVIDbaseline and follow up (tracked)] | 2 | 08 December 2020 |
| Other [RUDY-COVIDbaseline and follow up] | 2 | 08 December 2020 |
| Participant consent form [RUDY_Consent_Form_Blood_and_Urine_Collection] | 2 | 01 December 2020 |
| Participant consent form [RUDY_Consent_Form_Blood_and_Urine_Sample_Collection_V2(tracked)] | 2 | 01 December 2020 |
| Participant information sheet (PIS) [RUDY BLOOD AND URINE PIS Adult (tracked)] | 3 | 08 December 2020 |
| Research protocol or project proposal [RUDY Protocol V6 01 12 20] | 6 | 01 December 2020 |
| Research protocol or project proposal [RUDY_PROTOCOL_V6_01_12_20(tracked changes)] | 6 | 01 December 2020 |
| Sample diary card/patient card [RUDY Mitochondrial diary ] | 1 | 01 December 2020 |
| Validated questionnaire [Lubben short questionnaire] | 1 | 01 December 2020 |
| Validated questionnaire [SSQ] | 1 | 01 December 2020 |

**Membership of the Committee**

The members of the Committee who took part in the review are listed on the attached sheet.

**Working with NHS Care Organisations**

Sponsors should ensure that they notify the R&D office for the relevant NHS care organisation of this amendment in line with the terms detailed in the categorisation email issued by the lead nation for the study.

**Amendments related to COVID-19**

We will update your research summary for the above study on the research summaries section of our website. During this public health emergency, it is vital that everyone can promptly identify all relevant research related to COVID-19 that is taking place globally. If you have not already done so, please register your study on a public registry as soon as possible and provide the HRA with the registration detail, which will be posted alongside other information relating to your project.

**Statement of compliance**

The Committee is constituted in accordance with the Governance Arrangements for Research Ethics Committees and complies fully with the Standard Operating Procedures for Research Ethics Committees in the UK.

**HRA Learning**

We are pleased to welcome researchers and research staff to our HRA Learning Events and online learning opportunities– see details at: https://www.hra.nhs.uk/planning-and-improving-research/learning/

| **IRAS Project ID - 213780:** | **Please quote this number on all correspondence** |
|---|---|

Yours sincerely

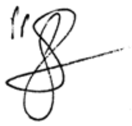

**Dr John Sheridan**
**Chair**

*Enclosures:* *List of names and professions of members who took part in the review*

*Copy to:* *Dr Muhammad K Javaid, University of Oxford*

### South Central - Berkshire B Research Ethics Committee

### Attendance at Sub-Committee of the REC meeting on 09 December 2020

#### Committee Members:

| *Name* | *Profession* | *Present* | *Notes* |
|---|---|---|---|
| Mrs Sue Harrison | Retired Managing Director of a Trade Association | Yes | |
| Dr John Sheridan | Consultant Toxicologist and Chemist | Yes | Meeting Chair |

#### Also in attendance:

| *Name* | *Position (or reason for attending)* |
|---|---|
| Ms Sarah Ferry | Approvals Administrator |
